# Local ancestry informed GWAS of warfarin dose requirement in African Americans identifies a novel CYP2C19 splice QTL

**DOI:** 10.1101/2025.03.03.25323247

**Authors:** Anmol Singh, Cristina Alarcon, Edith A Nutescu, Travis J. O’Brien, Matthew Tuck, Li Gong, Teri E. Klein, David O Meltzer, Julie A. Johnson, Larisa H Cavallari, Minoli A Perera

**Author notes:** Corresponding Author: Minoli A Perera (email address).

## Abstract

African Americans (AAs) are underrepresented in pharmacogenomics which has led to a significant gap in knowledge. AAs are admixed and can inherit specific loci from either their African or European ancestor, known as local ancestry (LA). A previous study in AAs identified single nucleotide polymorphisms (SNPs) located in the *CYP2C* cluster that are associated with warfarin dose. However, LA was not considered in this study. An IWPC cohort (N=340) was used to determine the LA-adjusted association with warfarin dose. Ancestry-specific GWAS’s were conducted with TRACTOR and ancestry tracts were meta-analyzed using METAL. We replicated top associations in the independent ACCOuNT cohort of AAs (N=309) and validated associations in a warfarin pharmacokinetic study in AAs. To elucidate functional roles of top associations, we performed short-read RNA-sequencing from AA hepatocytes carrying each genotype for expression of *CYP2C9* and *CYP2C19*. We identified 6 novel genome-wide significant SNPs (P<5E-8) in the CYP2C locus (lead SNP, rs7906871 (P=3.14E-8)). These associations were replicated (P≤2.76E-5) and validated with a pharmacokinetic association for S-Warfarin concentration in plasma (P=0.048). rs7906871 explains 6.0% of the variability in warfarin dose in AAs. Multivariate regression including rs7906871, previously associated SNPs, clinical and demographic factors explain 37% of dose variability, greater than previously reported studies in AAs. RNA-seq data in AA hepatocytes identified a significant alternate exon inclusion event between exons 6 and 7 in *CYP2C19* for carriers of rs7906871. In conclusion, we have found and replicated a novel CYP2C variant associated with warfarin dose requirement and potential functional consequences to C*YP2C19*.

## Introduction

African Americans (AAs) have been historically underrepresented in genetic studies. In published GWASs only 19% of individuals are non-European and less than 5% are non-European and non-Asian [1]. Furthermore, less than 10% of published GWASs focus on pharmacogenomics [2]. Combined, these two gaps in genomic research have created a lack of knowledge regarding the genetic association to drug response in AAs.

As AAs are an admixed population, they can have population-specific variants as well as allele frequency differences for shared variants. Consequences of these genomic differences in AAs have been studied across phenotypes [3]. In drug phenotypes, several studies have shown major differences between AAs and Europeans due to differences in genetic variation [4, 5]. For example, one study identified four African-specific SNPs located in chromosome 6 are associated with warfarin-related bleeding in AAs [4]. Another study found that rs28450894, a variant found predominantly in African populations, is significantly associated with albuterol response and was not found in prior studies done in Europeans [6]. Others have shown that AAs are at greater risk for adverse events from anticoagulants, including warfarin, and have worse outcomes when using traditional genotype-guided dosing based on studies of European ancestry [7].

AAs, due to their admixture, can inherit specific loci from either their African or European ancestor which is known as local ancestry (LA) [8]. This loci-specific variation in ancestry has been shown to play a role in drug-related phenotypes in AAs [9–11]. Several studies have incorporated LA into GWAS and expression quantitative trait loci (eQTL) mapping to identify novel results [8, 12, 13]. Specifically, a study by Yang et al (2022) found that LA plays a role in modulating clopidogrel response in AAs [9]. This study found that LA at the transcription start site (TSS) of *IRS-1*, *ABCB1*, and *KDR* were associated with platelet reactivity units (PRU), a measure of clopidogrel response, and identified variants located within *ITGA2* that are associated with increased PRU that were only found with LA adjustment [9]. These findings indicate that LA may be an important consideration in association studies of drug responses in admixed populations. Based on this, we aim to identify novel variants associated with warfarin dose requirement through incorporation of LA into a GWAS using the AA cohort previously published by Perera et al. (2013) [5]. Our findings were then replicated in an independent cohort as well as functionally validated. These analyses will help us to identify variants whose effect on warfarin dose are important to the dosing of warfarin in AAs.

## Materials and Methods

### GWAS Cohorts

We used a cohort of 345 AAs and 1,259 Europeans from various sites of the International Warfarin Pharmacogenomics Consortium (IWPC) [14] as listed in the supplemental methods (Supplemental Methods Tables 1 and 3) who were on a stable dose of warfarin, defined as having an International Normalized Ratio (INR) between 2 and 3 for three consecutive measures. Clinical and demographic measures are listed in the supplemental methods (Supplemental Methods Table 2). Five AA patients were removed from the analysis due to missing warfarin dose data, leaving 340 AA patients to be used as the discovery cohort for the analysis. Genotypes were re-imputed with 1000 Genomes Phase 3 [15] using the Michigan Imputation Server [16] and phased using Beagle version 5.4 [17] with the GRCh38 genetic map file from HapMap [18]. Standard pre-imputation and post-imputation quality control (QC) were performed as described in the supplemental methods. After QC, 6,778,974 SNPs were retained in the analysis. No subjects were removed due to relatedness, data missingness or sample QC. LA inference was done for the AA cohorts with FLARE [19] using the 1000 Genomes Phase 3 [15] YRI and CEU populations as ancestral reference populations.

LA inference, pre- and post-imputation QC, and TRACTOR analyses were run in the replication cohort from ACCOuNT [20] using the same procedure as the IWPC cohort. The ACCOuNT cohort consists of 309 AA patients on a stable dose of warfarin (Supplemental Methods Table 4) and clinical/demographic variables are listed in the supplemental methods (Supplemental Methods Table 5). All significant associations found in the discovery analysis were investigated in ACCOuNT. A p-value of 1E-3 for each ancestry tract and the meta-analysis was pre-specified as significant for replication.

Participants of both cohorts gave written informed consent to participate, and all protocols were approved by local institutional review boards.

### QC on Phenotypes and Covariates

All covariates previously shown to be associated to warfarin dose were tested for association. Warfarin dose was standardized by using a log2 transformation for all patients. The correlation between each of the variables and warfarin dose was tested by using a Pearson’s correlation test to determine the covariates to be included in the TRACTOR [12] GWAS.

### Genomic Analysis

#### TRACTOR Analysis

Using the LA estimates from FLARE [19], the genotypes, warfarin dose requirement, and covariates from the IWPC cohort as input we ran TRACTOR [12]. We specified the number of ancestries (--num-ancs) as two. A meta-analysis was done to combine the African-specific and European-specific estimates to get a LA-adjusted estimate for each SNP using METAL [21]. A p-value of 5E-8, 5E-6, and 1E-4 for each ancestry tract and the meta-analysis was pre-specified as genome-wide significant, suggestively significant, and nominally significant respectively.

We performed conditional analyses, adjusting for SNPs with known effects to warfarin dosing, including rs12777823, rs9923231 located at the *VKORC1* promotor and *CYP2C9* (*2, *3, *8, and *11 variants) (Table 1). *CYP2C9*\*5 and *6 could not be used due to their low minor allele frequency (MAF) in African populations. We performed a conditional analysis adjusting for *CYP2C9* (*2 and *3 variants) and rs9923231 using PLINK version 1.9 [22] in the IWPC European cohort (N=1,259) to assess if there are any ancestry-specific differences in the association of our top TRACTOR GWAS association from the AA analysis. *CYP2C9*\*8 and *CYP2C9*\*11 were excluded as these SNPs are rare in European populations.

**Table 1:**
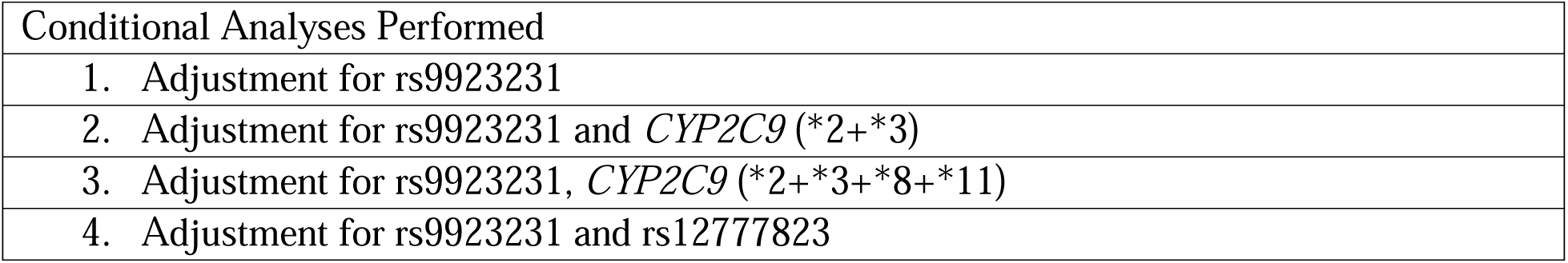
A list of the conditional TRACTOR analyses performed in the IWPC AA cohort. Each analysis was performed independently of each other and was compared to the non-conditioned TRACTOR analysis to assess if there were any changes in the significance of lead signals due to conditioning.

#### Percent of Variance in Warfarin Dose Explained

A percent of variance explained analysis was used to quantify how much variability in warfarin dose requirement a SNP contributes, indicating how much effect a SNP has on warfarin dose compared to other variables such as age, height, or other SNPs [23–25]. We first performed a univariate linear regression for age, height, weight, *CYP2C9*\*2, *CYP2C9*\*3, *CYP2C9*\*8, *CYP2C9*\*11, rs9923231, rs12777823, and rs7906871, on stable warfarin dose and calculated the r^2^ value for each regression to assess how much each variable tested individually contributes to the variance in stable warfarin dose for the IWPC European and IWPC AA cohorts. We then used stepAIC from the R package MASS [26] to perform a stepwise multivariate regression. This function utilizes the Akaike information criterion (AIC) to estimate the amount of information a variable contributes to the model by comparing a model that includes the variable to a model that does not include the variable. The model with the lowest AIC is considered optimal. The percent of variance each variable explains was judged by the overall r^2^ value for the multivariate model, in IWPC Europeans and AAs.

#### Functional Analysis

After the TRACTOR analyses, we performed functional fine mapping using FUMA [27]. FUMA prioritized top hits using liver and blood annotations from ENCODE [28] and Roadmap Epigenomics [29] which included histone marks, transcription factors, and heterochromatin annotations as well as assessing linkage disequilibrium (LD) between SNPs to identify independent risk loci. As input to FUMA we predefined a maximum p-value of lead SNPs as 5E-6, a maximum p-value cutoff of 0.05, a r^2^ threshold of 0.8, a MAF threshold of 0.01, LD block size of 250 kb, and 1000 Genomes Phase 3 [15] AFR populations as the reference panel. FUMA was also used eQTL mapped in blood and liver samples for our SNPs to assess the association of lead SNPs to gene expression in relevant tissues. The eQTL analysis was done through FUMA utilizing information from GTEx v8 [30], eQTLGEN [31], BloodeQTL[32], and BIOSQTL[33]. Finally, FUMA used chromatin interaction mapping to assess if any of the lead SNPs in our analysis overlap with any significant chromatin interaction regions in the liver. GTEx [30] v8 liver tissue was queried to identify any eQTL or sQTL associations to the top hits from the TRACTOR GWAS.

#### Genetic Analysis of Warfarin Pharmacokinetics

To analyze the association between lead SNPs to warfarin pharmacokinetic parameters, we performed genetic association analyses for our lead variant on multiple warfarin metabolites using pharmacokinetic data from 61 AA patients on warfarin as previously described [34]. The pharmacokinetic parameters in this dataset include ratio of steady state plasma concentration of R to S-enantiomer (RSCp), oral clearance of S-enantiomer (SCl), and the steady state plasma concentration of the S-enantiomer of warfarin (SCp). We performed separate analyses for each pharmacokinetic parameter and used a linear regression with age, sex, and PCs as covariates. Our results were compared to a warfarin pharmacokinetic GWAS done in 548 Sub-Saharan Africans by Asiimwe et al (2022) [35] to assess agreement.

### Alternative Splicing Analysis

Paired-end short-read RNA sequencing was performed on hepatocytes derived from AA donors carrying each genotype for expression of *CYP2C9* and *CYP2C19* (3 per genotype). The hepatocytes were purchased from commercial companies or isolated from cadaveric livers obtained from a collaboration with Gift of Hope. Hepatocytes were isolated from the cadaveric livers using the same procedure that was detailed in Park et al. (2019) [36]. Cells were plated at a density of 0.6 × 10^6^ cells/well in CP media (BioIVT) in collagen-coated plates with matrigel (Corning, Bedford, MA) overlay and incubated overnight at 37 °C. Cultures were maintained in HI media (BioIVT) supplemented with Torpedo antibiotic mix (BioIVT). RNA was isolated from liver tissue using the Qiagen miRNeasy mini kit (Cat #217004). The isolated RNA was checked for quality by measuring RNA concentration with NanoDrop and the RIN number from Bioanalyzer. Library preparation and RNA sequencing were then performed by Novogen (UC Davis, Sacramento, CA) using the NovaSeq PE150 sequencing platform. The resulting fastq files were analyzed for quality control using fastqc [37]. After quality control, STAR [38] was used to align the fastq files to the Homo sapiens Genome assembly GRCh38.p14 and to generate sorted bam files. Samtools [39] was used on the sorted bam files to filter low quality alignments (MAPQ<30) and duplicates were removed using Picard [40]. The bam files were then inputted into rMATS turbo version 4.3.0 [41] as well as the GENCODE v46 gene annotation file [42] to statistically quantify any alternative splicing events that occur within *CYP2C9* and *CYP2C19* based on genotype. As rMATS does a two-way comparison, we compared homozygous risk carriers to non-carriers as well as heterozygotes to non-carriers. We visualized alternative splicing events through sashimi plots [43] which show the reads contributing to the event versus the reads that do not contribute to the event. Percent of reads spliced in (PSI), a ratio of the reads that support the event’s inclusion over the total number of reads, between genotypes were calculated by rMATS.

## Results

### IWPC AA and ACCOuNT Cohorts

The IWPC AA cohort consists of 340 AAs and the ACCOuNT cohort consists of 309 AAs on a stable dose of warfarin (Table 2). Both cohorts clustered between the 1000 Genomes Phase 3 CEU and YRI samples as expected (Supplemental Figure 1). Age, weight, height, aspirin use, and amiodarone use all had a significant correlation with stable warfarin dose (Supplemental Table 1) and thus were used as covariates in the TRACTOR GWAS.

**Table 2:**
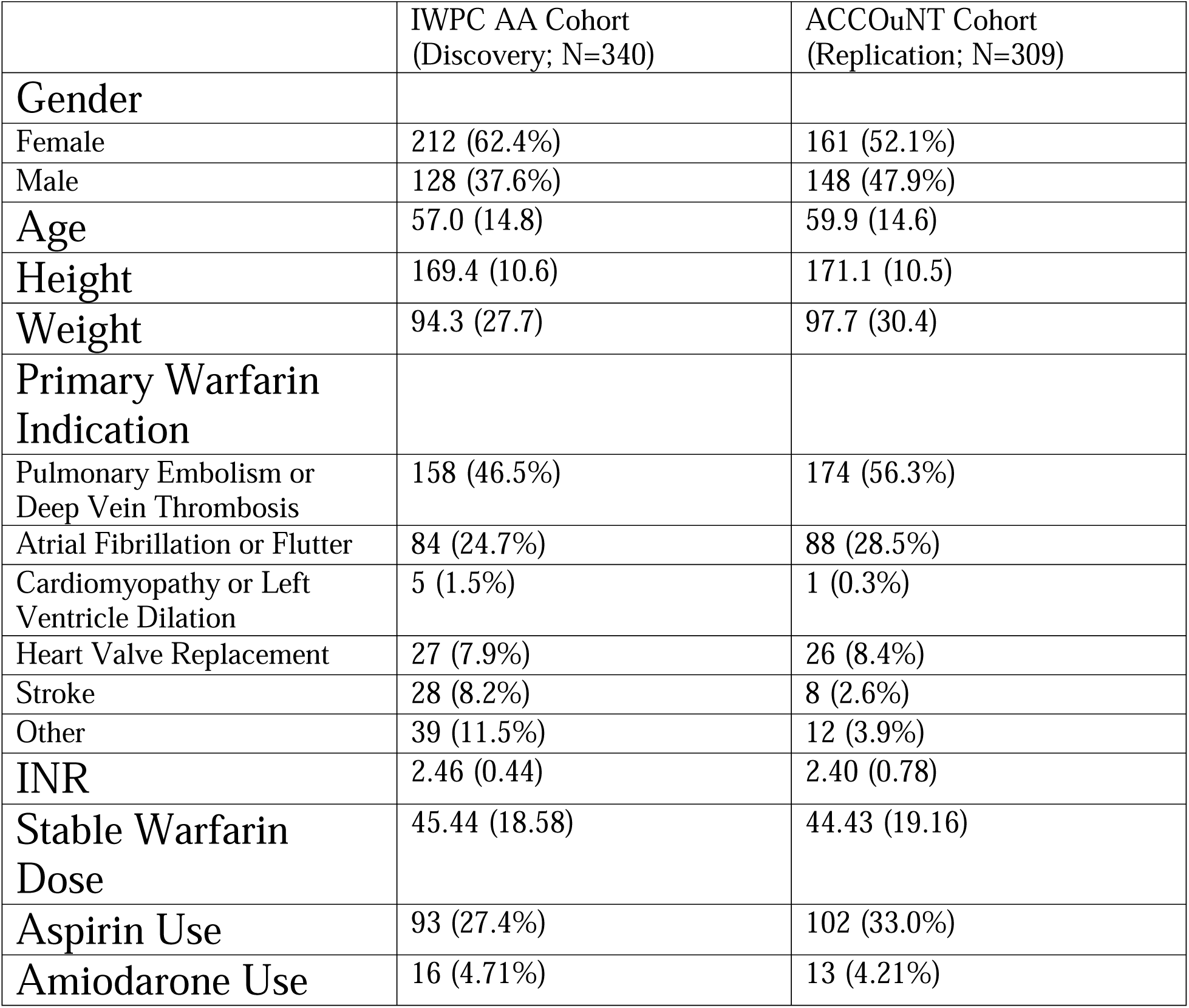
Clinical and Demographic characteristics of the discovery and replication cohorts. Values indicate either the number of participants (%) or mean (standard deviation).

|  | IWPC AA Cohort<br>(Discovery; N=340) | ACCOuNT Cohort<br>(Replication; N=309) |
| --- | --- | --- |
| **Gender** |  |  |
| Female | 212 (62.4%) | 161 (52.1%) |
| Male | 128 (37.6%) | 148 (47.9%) |
| **Age** | 57.0 (14.8) | 59.9 (14.6) |
| **Height** | 169.4 (10.6) | 171.1 (10.5) |
| **Weight** | 94.3 (27.7) | 97.7 (30.4) |
| **Primary Warfarin Indication** |  |  |
| Pulmonary Embolism or Deep Vein Thrombosis | 158 (46.5%) | 174 (56.3%) |
| Atrial Fibrillation or Flutter | 84 (24.7%) | 88 (28.5%) |
| Cardiomyopathy or Left Ventricle Dilation | 5 (1.5%) | 1 (0.3%) |
| Heart Valve Replacement | 27 (7.9%) | 26 (8.4%) |
| Stroke | 28 (8.2%) | 8 (2.6%) |
| Other | 39 (11.5%) | 12 (3.9%) |
| **INR** | 2.46 (0.44) | 2.40 (0.78) |
| **Stable Warfarin Dose** | 45.44 (18.58) | 44.43 (19.16) |
| **Aspirin Use** | 93 (27.4%) | 102 (33.0%) |
| **Amiodarone Use** | 16 (4.71%) | 13 (4.21%) |

### TRACTOR GWAS Identifies Novel Top Signal for Warfarin Dose

The TRACTOR African tract results revealed 128 variants that reach suggestive significance (P<5E-6) and are specific to African ancestry (Supplemental Table 2). No variant reached the pre-specified genome-wide significance in the African tract. Power was reduced in the European tract due to the low percentage of European ancestry in our cohort. For the meta-analysis combining the African and European tracts, we identified 6 genome-wide significant variants for warfarin dose and 49 variants that reach the suggestive significance threshold distinct from the findings in Perera et al (2013) [5] (Supplemental Table 4). The top signal from the meta-analysis was rs7906871 (P=3.14E-8), located between *CYP2C9* and *CYP2C19* (Figure 1). A meta-analysis of the LA-adjusted GWASs from the IWPC AA and ACCOuNT cohorts also found rs7906871 as a top signal (P=3.9E-12). Of the 49 SNPs that reached the suggestive significance threshold from the meta-analysis in the IWPC cohort, 35 were replicated in the ACCOuNT cohort with the same direction of effect (Supplemental Table 3) with the SNPs under P<1E-7 listed below (Table 3). Interestingly, previously identified warfarin dose associations lost power when LA was considered. The lead SNP from Perera et al (2013) [5], rs12777823, achieved a p-value of 2.2E-6 in the African tract and 8.3E-6 in the meta-analysis. The *VKORC1* variant, rs9923231, achieved a p-value of 2E-3 in the African tract and 4E-3 in the meta-analysis.

**Figure 1:**
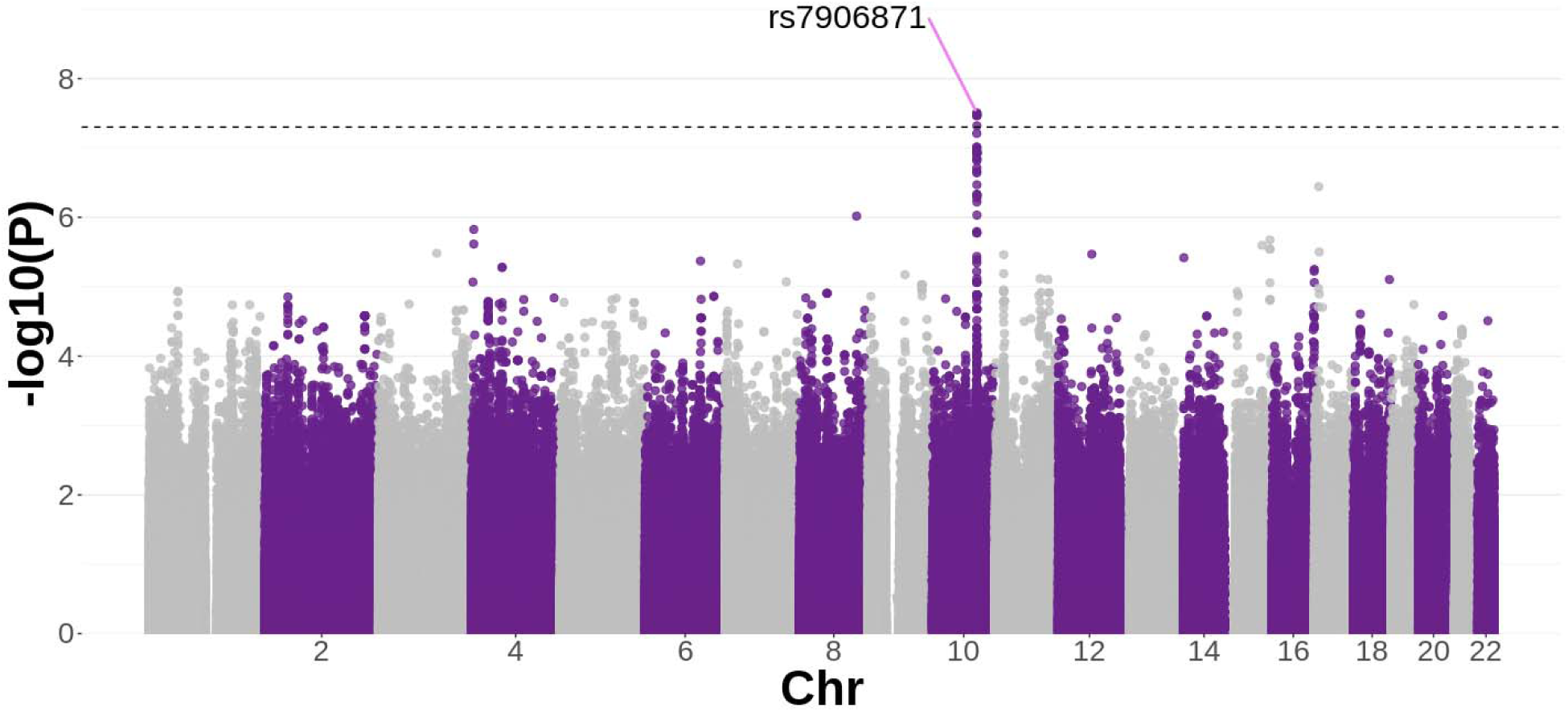
Manhattan Plot from the TRACTOR META Analysis. X-axis is the chromosome number and y-axis is the –log10(p-value) for each SNP from the TRACTOR META analysis. The colors of the points alternate to indicate odd (light grey) and even (purple) numbered chromosomes. The dashed line indicates the level for genome-wide significance (P=5E-8, -log10(P) = 7.301). The TRACTOR META GWAS identified a novel top signal for warfarin dose within the *CYP2C* Gene locus, rs7906871 (P=3.14E-8).

**Table 3:** Table of Top Signals that reached a significance threshold of P<1E-7 in the TRACTOR META analysis in the Discovery Cohort. All these top signals were replicated in the replication cohort with the same direction of effect which adds validity to these SNPs’ associations to warfarin dose requirement. *hg38 coordinates

| SNP | CHR | POS* | MAF (IWPC) | Beta (IWPC) | p-Value (IWPC) | MAF (ACCOuNT) | Beta (ACCOuNT) | p-Value (ACCOuNT) |
| --- | --- | --- | --- | --- | --- | --- | --- | --- |
| rs35013847 | 10 | 94892563 | 0.35 | -0.2378 | 3.14E-08 | 0.31 | -0.2293 | 3.67E-05 |
| rs7906871 | 10 | 94898443 | 0.35 | -0.2378 | 3.14E-08 | 0.31 | -0.2325 | 2.76E-05 |
| rs2104162 | 10 | 94876348 | 0.34 | -0.2374 | 3.41E-08 | 0.31 | -0.2325 | 2.76E-05 |
| rs4608002 | 10 | 94880345 | 0.34 | -0.2374 | 3.41E-08 | 0.31 | -0.2325 | 2.76E-05 |
| rs11596236 | 10 | 94912618 | 0.35 | -0.2383 | 3.43E-08 | 0.31 | -0.2325 | 2.76E-05 |
| rs34234478 | 10 | 94900915 | 0.35 | -0.2359 | 4.79E-08 | 0.31 | -0.2325 | 2.76E-05 |
| rs34367065 | 10 | 94726496 | 0.24 | -0.248 | 6.17E-08 | 0.19 | -0.242 | 2.16E-05 |
| rs1432019658 | 10 | 94997546 | 0.20 | -0.2476 | 7.76E-08 | 0.20 | -0.2087 | 0.0005238 |
| rs76413136 | 10 | 94691157 | 0.24 | -0.2412 | 9.71E-08 | 0.19 | -0.2539 | 6.72E-06 |
| rs78590583 | 10 | 94691572 | 0.24 | -0.2412 | 9.71E-08 | 0.19 | -0.2539 | 6.72E-06 |

Four distinct LD blocks in the IWPC AA cohort within the region encompassing rs7906871 and rs12777823 were found (Figure 2). rs12777823 is located in a separate LD block from rs7906871 suggesting that these two signals act independently of each other (r^2^=0.59). Furthermore, rs7906871 is not in LD with known *CYP2C* coding variants such as *CYP2C9*\*2 (r^2^=0.05), *CYP2C9*\*3 (r^2^=0.01), *CYP2C9*\*8 (r^2^=0.09), *CYP2C9*\*11 (r^2^=0.02), and *CYP2C19*\*2 (r^2^=0.48). We tested the independence of rs7906871 from known warfarin dose variants by performing several conditional TRACTOR GWAS analyses (Table 1). In these analyses, rs7906871 remained nominally significant for warfarin dose requirement (Supplemental Tables 5-7) even with adjustment of rs9923231, *CYP2C9*\*2, and *CYP2C9*\*3 (P=3.35E-7) as well as rs9923231, *CYP2C9*\*2, *CYP2C9*\*3, *CYP2C9*\*8, and *CYP2C9*\*11 (P=9.48E-5) indicating that the effect of this SNP is independent of these known *CYP2C* star alleles and rs9923231. However, we found that the signal for rs7906871 was lost when it was adjusted for rs9923231 and rs12777823 (P=0.77) indicating that there may be some interaction between rs12777823 and rs7906871 even though LD is relatively low between these SNP pairs. QQ plots for all GWASs done in the AA cohort are in the supplemental figures (Supplemental Figure 2). We did a conditional analysis in the IWPC European cohort to assess differences in the effect of these SNPs in a European cohort. rs7906871 is suggestively significant in the European cohort without conditioning (P=4.68E-6; Supplemental Table 8) and nominally significant when conditioned on just rs9923231 (P=1.43E-5; Supplemental Table 9). However, when conditioned on *CYP2C9* (*2 and *3 variants) and rs9923231, the association is no longer significant (P=0.12; Supplemental Table 10), indicating that in Europeans this SNP’s effect on warfarin dose is related to previously identified *CYP2C9* star alleles.

**Figure 2:**
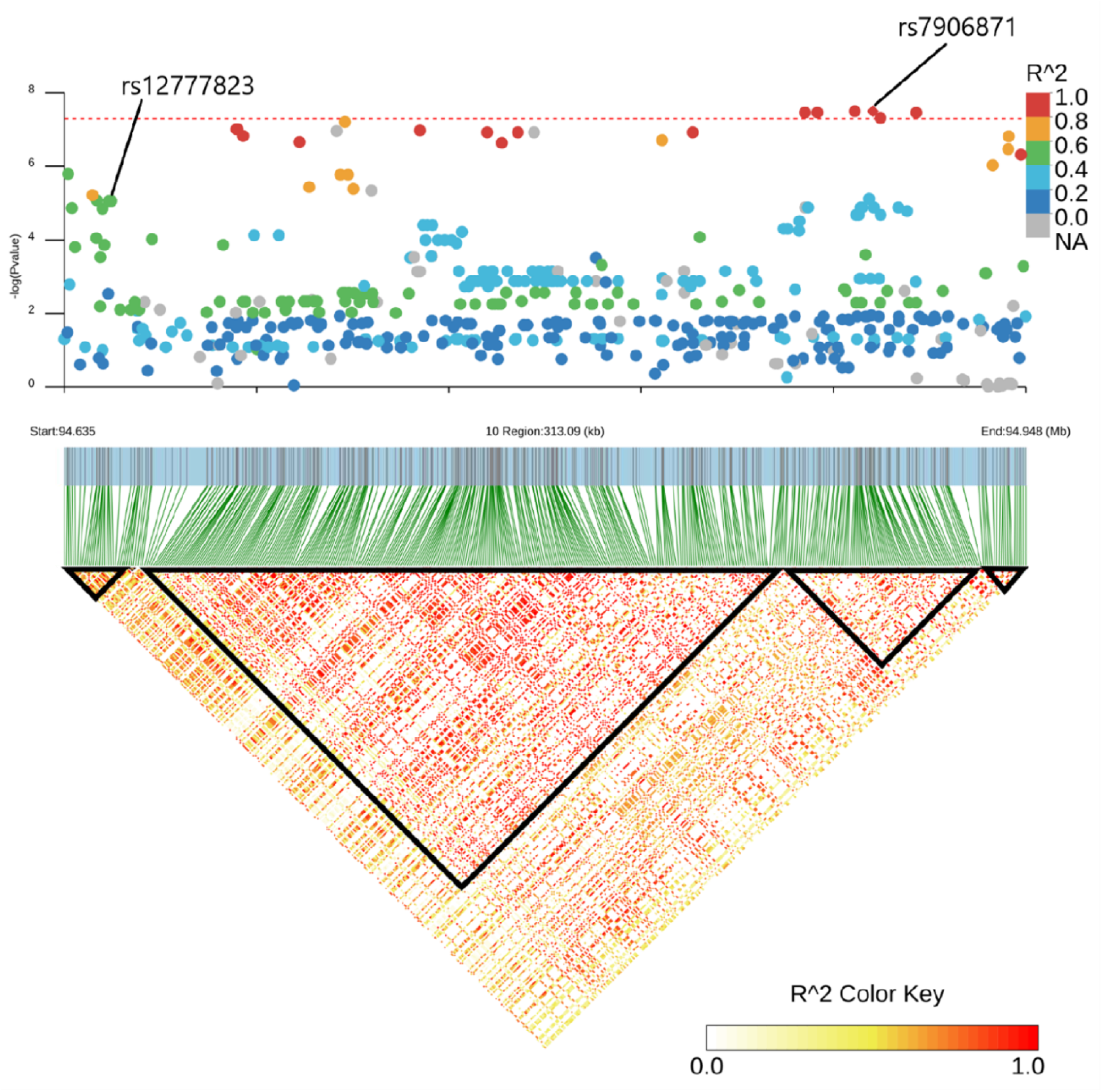
LD Blocks from LD analysis of region around rs7906871. This analysis revealed four distinct blocks of LD within this region (highlighted in the figure in black), one block contains rs12777823 and another contains rs7906871 which suggests that these two signals act independently of each other (r^2^=0.59). The dashed line indicates the threshold for Genome-Wide Significance (-log10(P)=7.3, P=5E-8). LD between all nearby SNPs and rs7906871 was calculated using the IWPC AA Cohort in Plink version 1.9 [22]. LD blocks were inferred using the LDBlockShow package [44].

### Percent of Warfarin Dose Variability Explained

In our univariate regressions on warfarin dose, we found that rs7906871 explains only 3.8% of warfarin dose variability in European populations. However, in AAs this SNP explains 6.0% of variability, similar to the 6.6% of variability explained by rs9923231 in AAs (Table 4). Furthermore, rs7906871 explains much more variability when compared to *CYP2C9* star alleles and rs12777823 in AAs (Table 4) indicating that this SNP contributes more to changes in warfarin dose requirement in AAs compared to these previously identified associations. In the stepwise multivariate regression done in IWPC AAs, rs9923231 (P=5.65E-8), *CYP2C9*\*3 (P=0.019), rs7906871 (P=2.75E-6), *CYP2C9*\*8 (P=7.18E-3), *CYP2C9*\*11 (P=0.036), age (P=1.21E-11), weight (P=2.74E-6), and height (P=0.073) were retained in the final model after AIC optimization with rs12777823 and *CYP2C9*\*2 excluded. Overall, the variables that contributed to the final multivariate model explained 37% of warfarin dose variability in AAs (Table 4). In the multivariate regression done in IWPC Europeans, rs9923231 (P<2.20E-16), CYP2C9*3 (P<2.20E-16), *CYP2C9*\*2 (P=2.75E-11), age (P<2.20E-16), weight (P=6.50E-4), and height (P=6.58E-8) were retained in the final model after AIC optimization with rs7906871 and rs12777823 excluded. Overall, the variables that contributed to the final multivariate model explained 35.6% of warfarin dose variability in Europeans (Table 4).

**Table 4:**
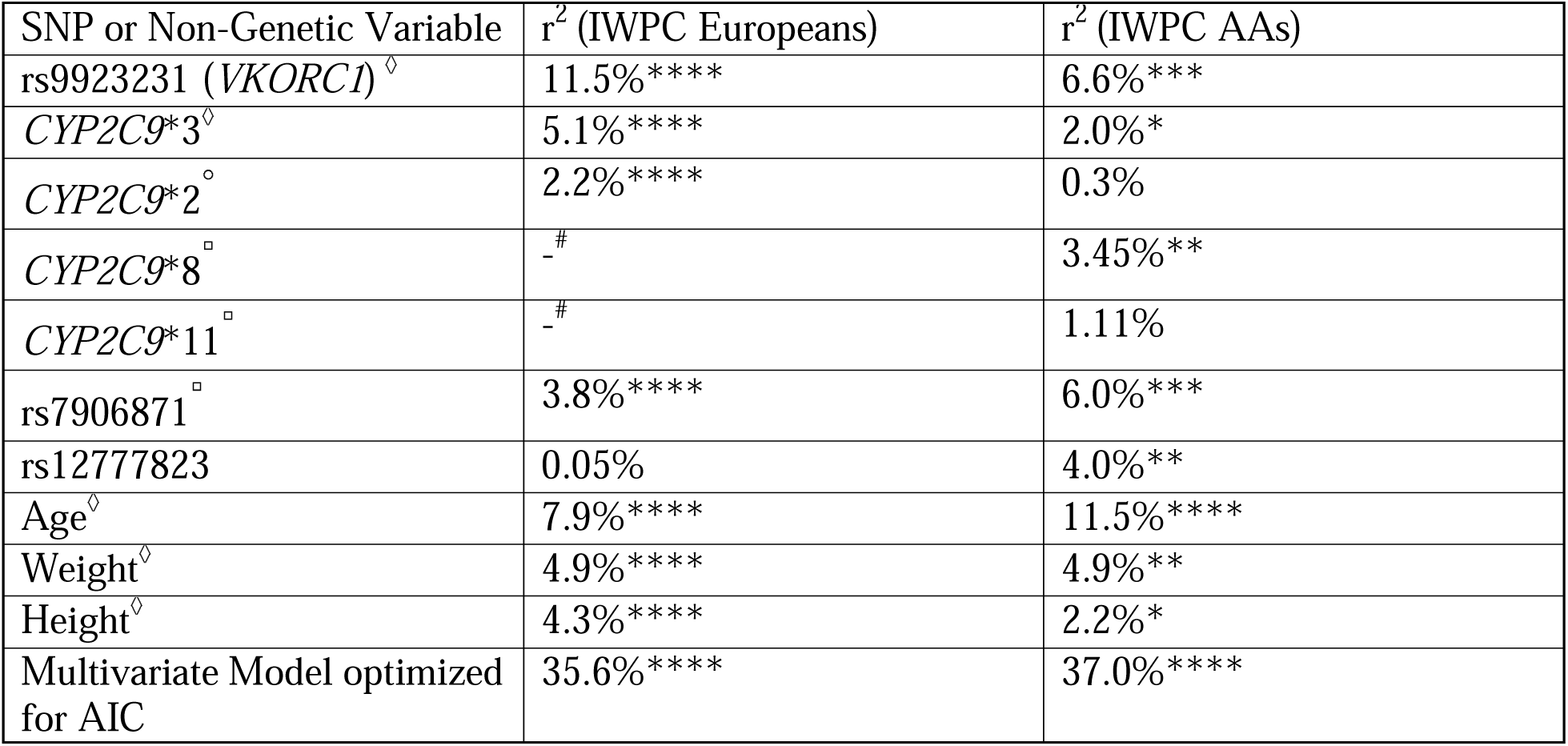
Table showing the percent of variability explained for key genetic variants, age, weight, and height for the IWPC European Cohort and the IWPC African American Cohort. Coefficient Estimate p-values from univariate analyses: **** P<1E-7, *** 1E-7≤ P<1E-5, ** 1E-5≤ P<1E-3, * 1E-3≤ P<0.05. Variables that were retained in the final multivariate model after AIC optimization in both populations are indicated by ◊ next to the variable name, in Europeans by ◦ next to the variable name, and in AAs by □ next to the variable name. ^#^ *CYP2C9*\*8 and *CYP2C9*\*11 were only included in AA cohort because their allele frequency is below 0.05% European populations.

| SNP or Non-Genetic Variable | $r^2$ (IWPC Europeans) | $r^2$ (IWPC AAs) |
| --- | --- | --- |
| rs9923231 (*VKORC1*)<sup>◇</sup> | 11.5% **** | 6.6% *** |
| *CYP2C9* *3<sup>◇</sup> | 5.1% **** | 2.0% * |
| *CYP2C9* *2<sup>◇</sup> | 2.2% **** | 0.3% |
| *CYP2C9* *8<sup>□</sup> | -# | 3.45% ** |
| *CYP2C9* *11<sup>□</sup> | -# | 1.11% |
| rs7906871<sup>□</sup> | 3.8% **** | 6.0% *** |
| rs12777823 | 0.05% | 4.0% ** |
| Age<sup>◇</sup> | 7.9% **** | 11.5% **** |
| Weight<sup>◇</sup> | 4.9% **** | 4.9% ** |
| Height<sup>◇</sup> | 4.3% **** | 2.2% * |
| Multivariate Model optimized for AIC | 35.6% **** | 37.0% **** |

### Functional Finemapping Results

Finemapping using FUMA revealed 13 independent genetic loci that have a significant association with ancestry-adjusted warfarin dose requirement (Supplemental Table 11). One specific locus in chromosome 10 had five independent lead SNPs (rs7906871, rs9332172, rs112835152, rs34367065, and rs12770901) with rs7906871 being the most significant SNP and the only one to reach genome-wide significance. Interestingly, rs12770901 while not reaching genome wide significance is in high LD with rs12777823 (r^2^=0.93). Another independent lead signal prioritized by FUMA, rs34367065, also has high LD with rs12777823 (r^2^=0.79) and reached close to genome-wide significance (P=6.17E-8). Chromatin interaction plots from FUMA revealed that rs7906871 is involved with 5 highly significant chromatin interactions within CYP2C locus (FDR = 4.52E-7 – 3.59E-23) in liver (Supplemental Figure 3).

### Genetic Association of Warfarin Pharmacokinetics (PK) measures reveal a significant association of rs7906871 to an increase in S-enantiomer concentration in plasma

To understand how rs7906871 impacts warfarin metabolism, we performed genetic association analyses on multiple warfarin PK measures from 61 AA patients on warfarin as previously described [34]. These analyses revealed that rs7906871 is significantly associated with a decrease in RSCp (β=-0.34, P=0.0028) as well as a significant increase in SCp (β=0.11, P=0.048).

### RNA-Sequencing reveals a significant increase in the inclusion of an alternative exon in *CYP2C19*

We queried GTEx v8 [30] and found that rs7906871 was a sQTL for *CYP2C19* in the liver (P=6.6E-7). Interestingly, no SNPs in our analysis were QTLs for *CYP2C9*, a gene previously found to be implicated with warfarin dose requirement across several populations.

From the short-read RNA sequencing data gathered from 9 hepatocytes (3 per genotype for rs7906871) derived from AA donors, we observed splicing differences in *CYP2C19* between exons 6 and 7 for patients carrying rs7906871. Specifically, we observed that patients who were heterozygous or homozygous for rs7906871 had a significant increase in the inclusion of an alternative exon between exons 6 and 7 (Figure 3A,B).

**Figure 3:**
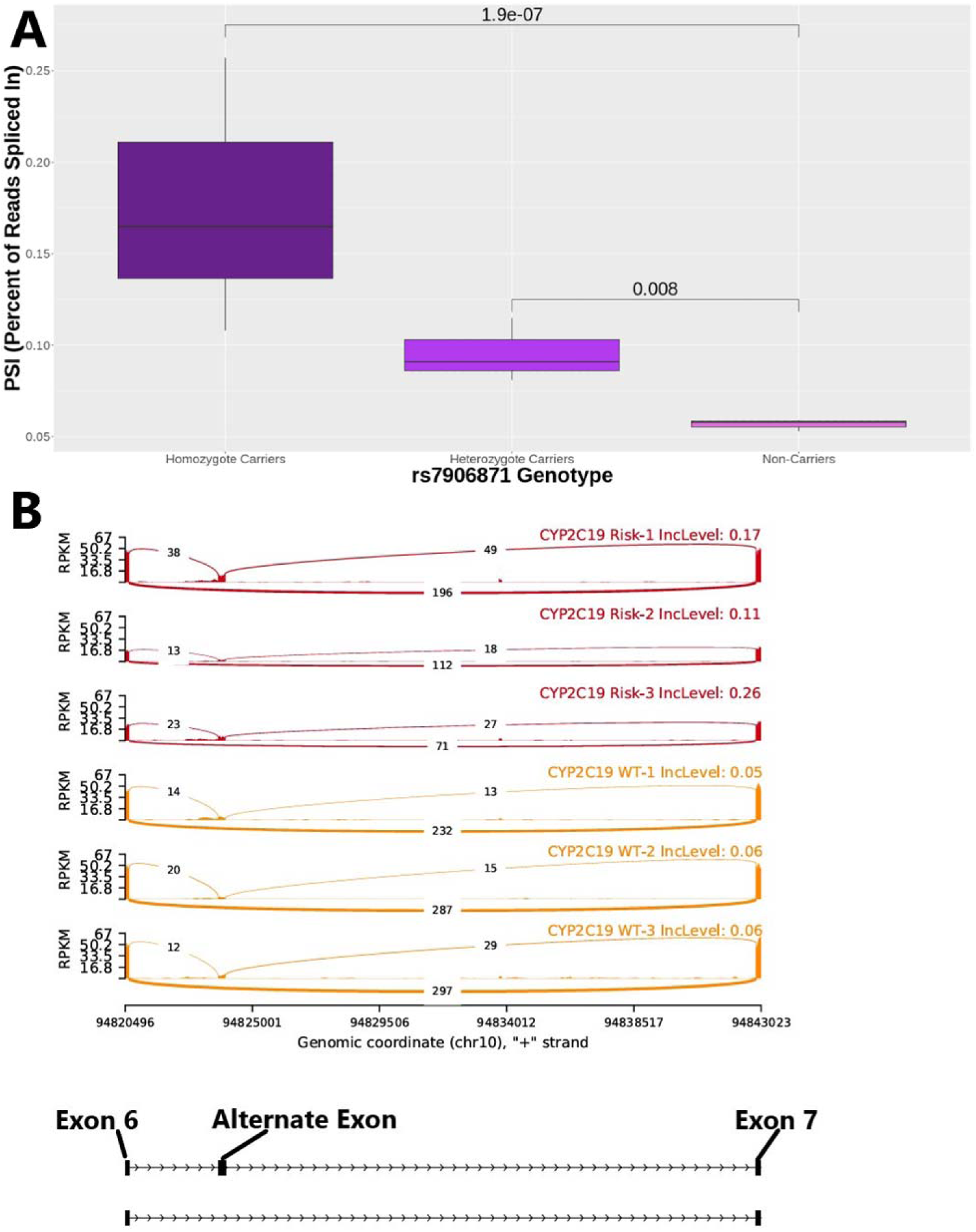
Results from alternative splicing analysis comparing rs7906871 genotype groups from AA hepatocytes for the inclusion of the alternative exon between exons 6 and 7. **(A)** Boxplots comparing the inclusion of the alternative exon for each genotype. Lines linking genotypes indicate the false discovery rate (FDR) from the comparisons of PSI for the alternative exon event. Percent Spliced In (PSI) is the ratio of the number of reads that contribute to the alternative exon inclusion event over the total number of reads, therefore a higher PSI indicates a higher inclusion of the alternative exon. Carriers of rs7906871 have a significant increase in the inclusion of an alternate exon located between exons 6 and 7 of *CYP2C19* using short-read RNA-sequencing data from AA hepatocytes. This comparison is significant for both homozygous carriers (FDR=1.9E-7) and Heterozygous carriers of rs7906871 (FDR=8E-3). **(B)** Sashimi plot from representative samples showing the inclusion of the alternative exon. Sashimi plots show the number of reads supporting the inclusion of the alternate exon between exons 6 and 7 (top reads of each panel) versus the number of reads supporting the exclusion of the alternate exon (bottom reads of each panel) for rs7906871 carriers (red samples) and non-carriers (yellow samples). The inclusion level shows the PSI for the alternative exon event for the representative sample. The x-axis of the plot shows the location on chromosome 10 and the y-axis shows the normalized read coverage or reads per kilobase per million mapped reads (RPKM). The panel below the x-axis shows the locations of Exon 6, the alternate exon, and Exon 7 of *CYP2C19* with the location corresponding to the position that the exons line up to on the x-axis. This figure shows that there are a higher proportion of reads supporting the inclusion of the alternate exon for rs7906871 carriers than in the non-carriers which indicates an increase in the inclusion of the alternate exon in rs7906871 carriers.

## Discussion

Using a LA-adjusted GWAS, we have identified novel genome-wide significant signals that are associated with warfarin dosing requirement in AAs. This signal at rs7906871 is independent of previously identified variants such as *CYP2C9* star alleles and rs9923231. We found an ancestry-specific effect of rs7906871 on warfarin dose requirement. When conditioned on the *CYP2C9* star alleles and rs9923231, the signal for this SNP is no longer significant (P=0.12) in the IWPC Europeans but remains nominally significant in IWPC AAs (P=9.4E-5). This may be due to the true causal allele being ancestry specific or the effect of LD differences in population allowing for the unmasking of this SNP’s effect. We did find, however, that conditioning on rs12777823 and rs9923231 resulted in a decreased significance of rs7906871 (P=0.77). This may be indicative of a relationship between rs12777823 and rs7906871. This may also indicate that rs7906871 is the SNP driving the association seen at rs12777823 in the previously published GWAS. Our variance-explained analysis also indicates there is a relationship between the two SNPs, with only rs7906871 retained in the final multivariate model and rs12777823 excluded. rs7906871 explains 6.0% of the variability in warfarin dose in AAs, which is greater than the variability explained by rs12777823 and *CYP2C9* star alleles in prior studies done in Europeans and AAs [23–25]. Our PK analysis showed that rs7906871 is significantly associated with a decrease in RSCp (β=-0.34, P=0.0028) as well as a significant increase in SCp (β=0.11, P=0.048).

There is no association to any PK parameters in Africans [35] or AAs with rs12777823. The S-enantiomer of warfarin is more potent than the R enantiomer and is responsible for producing warfarin’s anticoagulant effect [45]. This suggests that rs7906871 decreases S-warfarin metabolism leading to a lower warfarin dose requirement for carriers of the SNP. These findings are concordant with the direction of effect found from the TRACTOR GWAS. Asiimwe et al (2022) [35], a warfarin PK GWAS done in Sub-Saharan Africans, showed agreement with our findings with rs7906871 showing significant association with decreased RSCp (β=-0.28, P=6.6E- 7).

GTEx identified rs7906871 as a sQTL for *CYP2C19* in liver tissue and we found the SNP significantly increases the inclusion of an alternate exon located between exons 6 and 7 of *CYP2C19* in our short-read RNA sequencing data obtained from AA hepatocytes. This indicates that our top association may be involved in splicing changes within *CYP2C19* potentially related to changes in warfarin dose requirement. Previous studies have found only weak evidence linking *CYP2C19*2*, the main functional variant for *CYP2C19*, to warfarin dose requirement [46]. *CYP2C19* is involved in the R-enantiomer metabolism pathway and does not currently have a clear role in S-enantiomer metabolism [47]. This indicates that *CYP2C19* may play a lesser role in determining warfarin dose than *CYP2C9*, which metabolizes the S-enantiomer. Additionally, *CYP2C19* does not have a recognized role in warfarin pharmacogenomics as it is mainly variants within *VKORC1* and *CYP2C9* that have been used to guide warfarin dosing in the CPIC guidelines [48]. In this study, *CYP2C19*2* was not significantly associated with warfarin dose requirement. Additional studies are needed to fully elucidate the role of *CYP2C19* on warfarin metabolism.

Interestingly, the SNPs found to be associated with warfarin dose requirement using traditional GWAS methods in the same cohort decreased in significance substantially. For rs9923231, located within *VKORC1*, we saw a substantial decrease in significance with the addition of LA, with the SNP only achieving a p-value of 4E-3 in the meta-analysis and 2E-3 in the African tract of the TRACTOR GWAS. This same pattern for rs9923231 was seen in another study which conducted a TRACTOR GWAS in African and AA patients on warfarin [11]. This study also found a large increase in the p-value for rs9923231 in the meta-analysis of the African tracts of their cohorts, from P= 1.24E-15 in their traditional GWAS to a P = 1.6E-8 [11]. While rs9923231 is still genome-wide significant, the study included mixed-ancestry populations from sub-Saharan Africa who have different admixture patterns than AAs. Furthermore, when looking at the TRACTOR African tracts for only their AA cohorts, the p-value for rs9923231 is similar to what we find in our study (P=2.3E-2 – 1.5E-3) [11]. This may also be the result of European admixture present in AA that is absent in Africans, which may change the LD pattern uniquely in AAs. This difference in association at rs9923231 could also be due to the similar effect size of this SNP in European and African populations, leading to a decrease in the power to detect associations when using TRACTOR - a limitation that has been previously described [12, 49–51]. We noticed a similar trend for rs12777823 in our TRACTOR GWAS. This SNP occurs at common frequencies in AAs and Europeans but only has a significant effect on warfarin dosing in AA populations [5]. Therefore, we hypothesized that the significance would increase in the African tract of the TRACTOR analysis, which would confirm an African-specific effect for warfarin dosing for this SNP. Though the significance did decrease, the AFR tract p-value for rs12777823 is 2E-6 which still indicates suggestive significance.

One major limitation of this analysis is our sample size. Although we reached genome-wide significance for warfarin dose requirement, there may be some associations that were missed due to decreased power. Furthermore, due to this low sample size, we could not assess any European-ancestry specific associations as there was limited power to detect genetic associations in the European segments of this cohort. This limitation was somewhat mitigated by the incorporation of European ancestry segments into a meta-analysis. However, it would be interesting to analyze the European ancestry segments of the AA genome separately in a larger cohort. Our replication of rs7906871 in an independent AA cohort, as well as of this SNP’s association to warfarin metabolism in a PK cohort, adds robustness to our findings. The splicing analysis in AA hepatocytes provides causal evidence that this SNP has a functional effect on *CYP2C19*. However, larger long-read sequencing studies are needed to identify the true causal variant and the full functional effects on protein and enzyme function.

In summary, we have identified novel genetic variants associated with warfarin dosing requirement and a variant that is associated with alternative splicing in *CYP2C19* for AAs that was only found when taking LA into consideration. This finding underlies the importance of considering LA in genetic analyses involving AAs and the need for further dosing studies to be done in minority populations to personalize warfarin dosing regimens for these populations.

## Data Availability

Summary statistics data may be found in a data supplement available with the online version of this article and on FigShare. Genotype and phenotype data for the ACCOuNT cohort will be deposited to dbGAP at the time of publication.

## Acknowledgements

This work was supported by the National Institute of Diabetes and Digestive and Kidney Diseases (U2CDK129917 and TL1DK132769), the National Institute on Minority Health and Health Disparities (1U54MD010723-01), and the American Heart Association Midwest Affiliate Spring 2010 Grant-In-Aid (10GRNT3750024). We would like to thank Hyunyoung Jeong at Purdue University for providing data and expertise that was invaluable to this project.

## Author Contributions

AS and MAP conceived the project and wrote the manuscript. AS conducted the computational analyses and made the figures. CA ran the short-read RNA sequencing experiments. EAN, TJO, MT, LG, TEK, DOM, JAJ, and LHC contributed to the acquisition of ACCoUNT and IWPC data and helped with edits to the drafts of the manuscript. The Authors declare no competing financial interests.

## Notes

### Competing Interest Statement

The authors have declared no competing interest.

