## Supplemental Methods and Figures for "Local ancestry informed GWAS of warfarin dose requirement in African Americans identifies a novel CYP2C19 splice QTL"

**Cohorts**

**IWPC AA Cohort**

The IWPC AA cohort is a cohort of AAs on a stable dose of warfarin, defined as having an INR between 2 and 3 for 3 consecutive measures while on warfarin for at least 3 weeks. The cohort is comprised of patients from several different sites detailed below and consists of 345 total patients (Supplemental Methods Table 1). All IWPC AA patients were over the age of 18 and gave consent to participate in the study. Clinical and demographic factors that have been previously associated with stable warfarin dose were gathered from IWPC sites for all participants if they were available (Supplemental Methods Table 2).

| IWPC Site | Number of Patients |
| --- | --- |
| Vanderbilt University, Nashville, TN | 34 |
| University of Florida, Gainesville, FL | 21 |
| University of Illinois at Chicago, Chicago, IL | 122 |
| University of Chicago, Chicago, IL | 115 |
| University of California, San Francisco, CA | 6 |
| Aurora Healthcare, Downers Grove, IL | 34 |
| Stanford University, Stanford, CA | 3 |
| University of North Carolina, Chapell Hill, NC | 10 |

**Supplemental Methods Table 1: List of all IWPC Sites that IWPC AA participants originated from and number of IWPC AA patients that each site contributed.**

| **Clinical or Demographic Variable** | **Possible Values and Units** |
| --- | --- |
| Gender | Male, Female, or Not Known |
| Race | Self-Reported based on United States Census Categories |
| Ethnicity | Self-Reported based on United States Census Categories |
| Age | Reported as a binned age based on decade (0-9,10-19,20-29,30-39,40-49,50-59,60-69,70-79,80-89,90+) and units are years |
| Height | Reported in centimeters (cm) |
| Weight | Reported in kilograms (kg) |
| Indication for Warfarin Treatment | DVT, PE, Afib/flutter, Heart Valve, Cardiomyopathy/LV Dilation, Stroke, Post-Orthopedic, Other or Not Known |
| Comorbidities | List of diseases that are co-occurring with primary phenotype in patients (Yes/Not Present/Not Known are possible values for all diseases):   - Diabetes - Congestive Heart Failure and/or Cardiomyopathy - Valve Replacement |
| Medications | List of medications patient has taken or is currently taking (Yes/ Not Present/ Not Known are possible values for all medications):   - Aspirin - Acetaminophen - Simvastatin - Atorvastatin - Fluvastatin - Pravastatin - Rosuvastatin - Cerivastatin - Amiodarone - Carbamazepine - Phenytoin - Rifampin - Sulfonamide Antibiotics - Herbal Medications, Vitamins, and Supplements |
| Was Dose of  Acetaminophen  >1300mg/day | Yes or No are possible values |
| Target INR | Possible values are the value of the Target International Normalized Ratio or Not  Known  Reported as single number or  average of minimum and maximum in case of provided range. |
| Subject Reached Stable  Dose of Warfarin | Possible values are Yes, No, Not Known |
| INR on Reported  Therapeutic Dose of  Warfarin | Reported INR from IWPC Site for Patient when they are on the therapeutic dose of Warfarin |
| Therapeutic Dose of Warfarin | Reported in milligrams (mg) per week |
| Current Smoker | Possible values are yes, not present, not known |

**Supplemental Methods Table 2: Variables collected by the IWPC AA Cohort Sites.** List of Clinical and Demographic variables that have been shown to be related to stable warfarin dose in prior studies that have been collected by IWPC sites. Variable Names and the values/units reported are listed in the table.

**IWPC European Cohort**

The IWPC European cohort is a cohort of Europeans on a stable dose of warfarin, defined as having an INR between 2 and 3 for 3 consecutive measures while on warfarin for at least 3 weeks. The cohort is comprised of patients from several different sites detailed below and consists of 1,259 total patients (Supplemental Methods Table 3). All IWPC European patients were over the age of 18 and gave consent to participate in the study. Clinical and demographic factors that have been previously associated with stable warfarin dose were gathered from IWPC sites for all participants if they were available (Supplemental Methods Table 2).

| IWPC Site | Number of Patients |
| --- | --- |
| RIEDER, University of Washington, Seattle, WA | 176 |
| LIBWAR, University of Liverpool, Liverpool, UK | 327 |
| WARG, Uppsala University, Uppsala, Sweeden | 756 |

**Supplemental Methods Table 3: List of all IWPC Sites that IWPC European participants originated from and number of IWPC European patients that each site contributed.**

**ACCOuNT Cohort**

The ACCOuNT cohort consists of AAs on a stable dose of warfarin, defined as having an INR between 2 and 3 for 3 consecutive measures while on warfarin for at least 3 weeks. All patients are older than 18 years of age and were on warfarin for longer than 3 months. Participants were excluded from the study if they had a hemoglobin<7.0 g/dL at enrollment, life expectancy < 8 weeks, active cancer diagnosis, inability or unwillingness to comply with study protocols, history of significant bleeding that required hospitalization 6 months prior to the start of the study, contraindications to treatment with anticoagulants, platelet count < 100,000 mm^3^, history or current alcohol/drug abuse within 1 year of enrollment, history of hypersensitivity to warfarin, concurrent therapy with another antiplatelet (low doses of Aspirin and NSAIDs are permissible). The cohort is comprised of patients from several different sites detailed below and has 340 total patients (Supplemental Methods Table 4). 31 patients were excluded from the study for missing data. Clinical and demographic factors that have previously been associated with stable warfarin dose were gathered from ACCOuNT sites for all participants if they were available (Supplemental Methods Table 5).

| ACCOuNT Site | Number of Patients |
| --- | --- |
| Northwestern University, Chicago, IL | 63 |
| University of Illinois, Chicago, IL | 133 |
| University of Chicago, Chicago, IL | 58 |
| Veterans Affairs, Washington, DC | 65 |
| George Washington University, Washington, DC | 21 |

**Supplemental Methods Table 4: List of all ACCOuNT Sites that ACCOuNT Warfarin participants originated from and number of patients that each site contributed.**

| **Clinical or Demographic Variable** | **Possible Values and Units** |
| --- | --- |
| Gender | - M for Male - F for Female - MD for Missing Data |
| Age | Reported in Years |
| Height | Reported in Centimeters (cm) |
| Weight | Reported in Kilograms (kg) |
| List of Comorbidities | List of diseases that are co-occuring in patient |
| Has Diabetes | - 1 for Type 1 Diabetes - 2 for Type 2 Diabetes - 0 for Not Present - MD for Missing Data |
| Has Congestive Heart Failure or Cardiomyopathy | - Y for yes - N for no - MD for missing data |
| Has Valve Replacement | - Y for yes - N for no - MD for missing data |
| Has Hypertension | - Y for yes - N for no - MD for missing data |
| Has Hypercholesteremia | - Y for yes - N for no - MD for missing data |
| Is a Current Smoker | - Y for yes - N for no - MD for missing data |
| Is a Former Smoker | - Y for yes - N for no - MD for missing data |
| How Long Has Patient Been a Smoker | Reported in Years |
| Alcohol Use | - 0 for None - 1 for Rare, fewer than 12 drinks in the past year - 2 for Infrequent, 1 to 13 drinks per month - 3 for Moderate, 4 to 14 drinks per week - 4 for Frequent, more than 2 drinks per day - MD for Missing Data |
| Indication for Warfarin Treatment | - Deep Vein Thrombosis - Pulmonary Embolism - Heart Valve - Stroke - Afib/Flutter - Cardiomyopathy/ Left Ventricle Dialation - MD for Missing data |
| Therapeutic Warfarin Dose | Reported in milligrams/week (mg/wk) |
| Time on Treatment at Enrollment | Reported in days |
| Target INR | Possible values are the value of the Target International Normalized Ratio or MD for missing data |
| Was Stable Dose Reached | - Y for Yes - N for No - MD for Missing Data |
| INR on Therapeutic Dose of Warfarin | Reported INR from ACCOuNT Site for Patient when they are on the therapeutic dose of Warfarin |
| Aspirin Use | - Y for Yes - N for No - MD for Missing Data |
| Aspirin Daily Dose | Reported in milligrams/day (mg/day) |
| Clopidogrel Use | - Y for Yes - N for No - MD for Missing Data |
| Dipyridamole Use | - Y for Yes - N for No - MD for Missing Data |
| Acetaminophen or Paracetamol (Tylenol) | - Y for Yes - N for No - MD for Missing Data |
| Acetaminophen/Paracetamol Daily Dose | Reported in milligrams/day (mg/day) |
| NSAID Use | - Y for Yes - N for No - MD for Missing Data |
| NSAID Daily Dose | Reported in milligrams/day (mg/day) |
| Simvastatin (Zocor) Use | - Y for Yes - N for No - MD for Missing Data |
| Atorvastatin (Lipitor) Use | - Y for Yes - N for No - MD for Missing Data |
| Fluvastatin (Lescol) Use | - Y for Yes - N for No - MD for Missing Data |
| Lovastatin (Mevacor) Use | - Y for Yes - N for No - MD for Missing Data |
| Pravastatin (Pravachol) Use | - Y for Yes - N for No - MD for Missing Data |
| Rosuvastatin (Crestor) Use | - Y for Yes - N for No - MD for Missing Data |
| Cerivastatin (Baycol) Use | - Y for Yes - N for No - MD for Missing Data |
| Amiodarone (Cordarone) Use | - Y for Yes - N for No - MD for Missing Data |
| Carbamazepine (Tegretol) Use | - Y for Yes - N for No - MD for Missing Data |
| Phenytoin (Dilantin) Use | - Y for Yes - N for No - MD for Missing Data |
| Rifampin or Rifampicin Use | - Y for Yes - N for No - MD for Missing Data |
| Sulfonamide Antibiotics Use | - Y for Yes - N for No - MD for Missing Data |
| Macrolide Antibiotics Use | - Y for Yes - N for No - MD for Missing Data |
| Anti-fungal Azoles Use | - Y for Yes - N for No - MD for Missing Data |
| Herbal Medications, Vitamins, Supplements Use | - Y for Yes - N for No - MD for Missing Data |
| Bleeding Academic Research Consortium (BARC) bleeding score | - Type 0: No Bleed - Type 2: any overt, actionable sign of hemorrhage (eg, more bleeding than would be expected for a clinical circumstance, including bleeding found by imaging alone) that does not fit the criteria for type 3, 4, or 5 but does meet at least one of the following criteria: (1) requiring nonsurgical, medical intervention by a healthcare professional, (2) leading to hospitalization to increase level of care, or (3) prompt evaluation. - Type 3a: Overt bleeding plus hemoglobin drop of 3 to 5 g/dL* (provided hemoglobin drop is related to bleed), Any transfusion with overt bleeding - Type 3b: Overt bleeding plus hemoglobin drop greater than or equal to 5 g/dL (provided hemoglobin drop is related to bleed), Cardiac tamponade, Bleeding requiring surgical intervention for control (excluding dental/nasal/skin/hemorrhoid), Bleeding requiring intravenous vasoactive agents - Type 3c: Intracranial hemorrhage (does not include microbleeds or hemorrhagic transformation, does include intraspinal), Subcategories confirmed by autopsy or imaging or lumbar puncture, intraocular bleeding compromising vision. - Type 5: Fatal bleed. - Missing Data/not known = MD |
| Time to Bleeding Event | Calculated by Date of Bleeding Event – Enrollment Date (units are days) |
| INR at Bleeding Event | Reported INR from ACCOuNT Site for Patient at the time bleeding event occured |
| Embolic Event | - None if no events occured - DVT Deep Vein Thrombosis - PE for Pulmonary Embolism - Stroke - Myocardial Infarction - MD for Missing Data |
| Time to Embolic Event | Calculated by Date of Embolic Event – Enrollment Date (units are days) |
| INR at Embolic Event | Reported INR from ACCOuNT Site for Patient at the time embolic event occured |
| Patient Deceased? | - Y for Yes - N for No - MD for Missing Data |
| Time to Death | - Calculated by Date of Bleeding Event – Enrollment Date (units are days) - MD for Missing Data |
| eGFR < 30 ml/min/1.73m^2^ ? | - Chronic Kidney Disease is defined by eGFR < 30 ml/min/1.73m^2^ - Y for Yes - N for No - MD for Missing Data |
| Platelet Count | Reported in cells/microliter (cells/µL) |
| Hemaglobin | Reported in grams/deciliter (g/dL) |

**Supplemental Methods Table 5: Variables collected by the ACCOuNT Cohort Sites.** List of Clinical and Demographic variables that have been shown to be related to stable warfarin dose in prior studies that have been collected by ACCOuNT sites. Variable Names and the values/units reported are listed in the table.

**Pre-imputation QC**

Pre-imputation QC was performed prior to imputation with Beagle version 5.4 [1] for both the IWPC cohorts and the ACCOuNT cohort in the same way. First, we applied a filter on SNPs with a call rate below 0.95 and missingness greater than 0.05 and no SNPs were excluded from the analysis due to this threshold. Next, a minor allele frequency (MAF) threshold of 0.05 was applied so only SNPs with a MAF>0.05 were retained which removed 6,966,233 SNPs from the analysis. We excluded 311 SNPs that failed the Hardy-Weinberg Equilibrium (HWE) test (HWE_p-value_ < 1E-6). SNPs were quality checked for match with 1000 Genomes Phase 3 [2] as well as strand flip issues using the 1000 Genome imputation preparation tool suggested by the Michigan Imputation Server [3] (available at this link: <https://www.chg.ox.ac.uk/~wrayner/tools/>). From this tool, 218,458 SNPs had strand flips, and 381 SNPs were removed that did not match 1000 Genomes Phase 3 [2]. SNPs on X and Y chromosomes as well as mitochondrial SNPs were removed. Genome-wide genotypes for all patients were used to determine gender misspecification and identity by descent (IBD). No samples were removed due to gender misspecification, IBD<0.125, genotype call rate<0.95, or missingness>0.05.

**Post-Imputation QC**

We performed post-imputation QC for both the IWPC and ACCOuNT cohorts retaining 6,778,974 SNPs with an imputation quality score (INFO) > 0.8, minor allele frequency (MAF) >0.05, and Hardy-Weinberg_p-value_ >1E-6.

**Supplemental Figures**

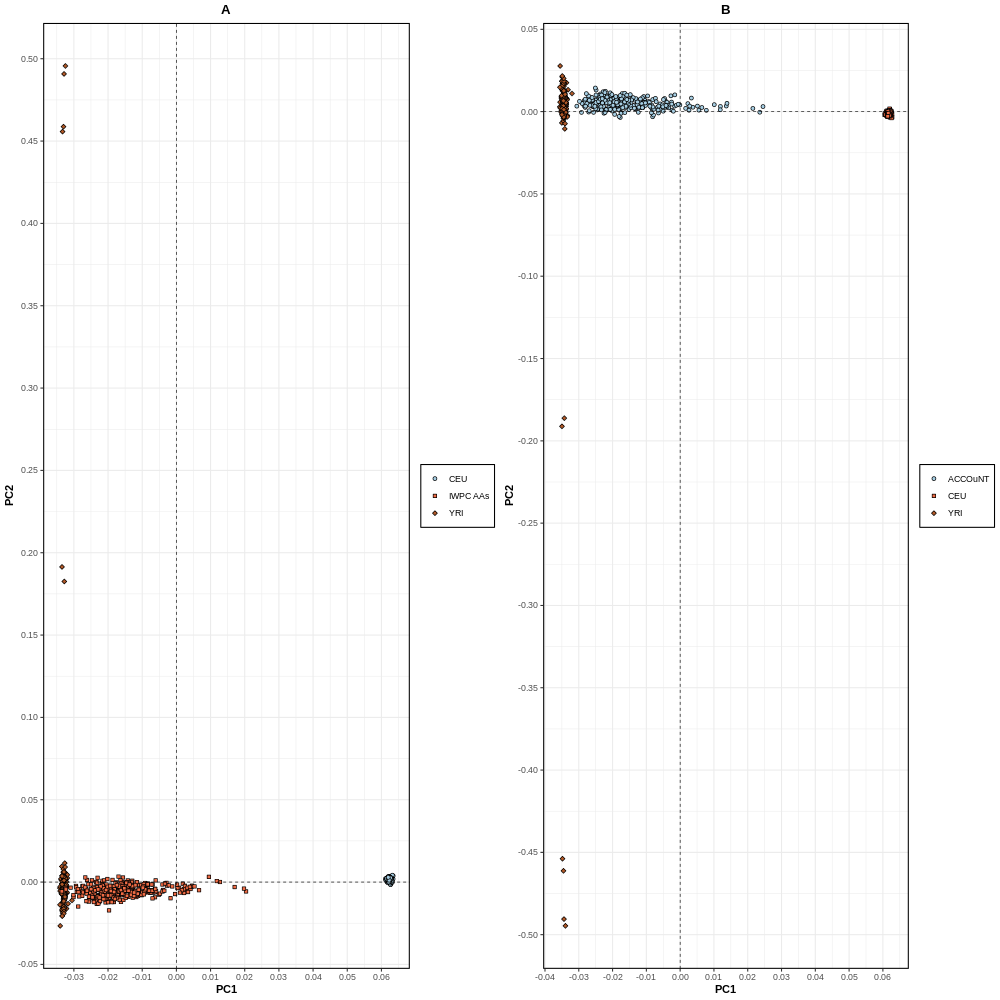

**Supplemental Figure 1. PC plots comparing IWPC AA cohort and ACCOuNT cohort to 1000 Genomes [2] YRI, and 1000 Genomes [2] CEU Reference Populations. A.** Comparison of IWPC AAs against 1000 Genomes CEU and YRI. Points on the plot represent each sample with the shape indicating the cohort that the sample is from: Diamond (YRI), Circle (CEU), and Square (IWPC AAs). **B.** Comparison of ACCOuNT cohort against 1000 Genomes CEU and YRI. Points on the plot represent each sample with the shape indicating the cohort that the sample is from: Diamond (YRI), Circle (ACCOuNT), and Square (CEU). Shows that there are distinct clusters of YRI and CEU with the admixed IWPC AA and ACCOuNT cohorts falling in the middle of these two parental populations, which is what we would expect. PCs were calculated using PLINK version 1.9 [4] and PC plots were made using pca_plot function from the gwaRs package [5] in R version 4.4.0.

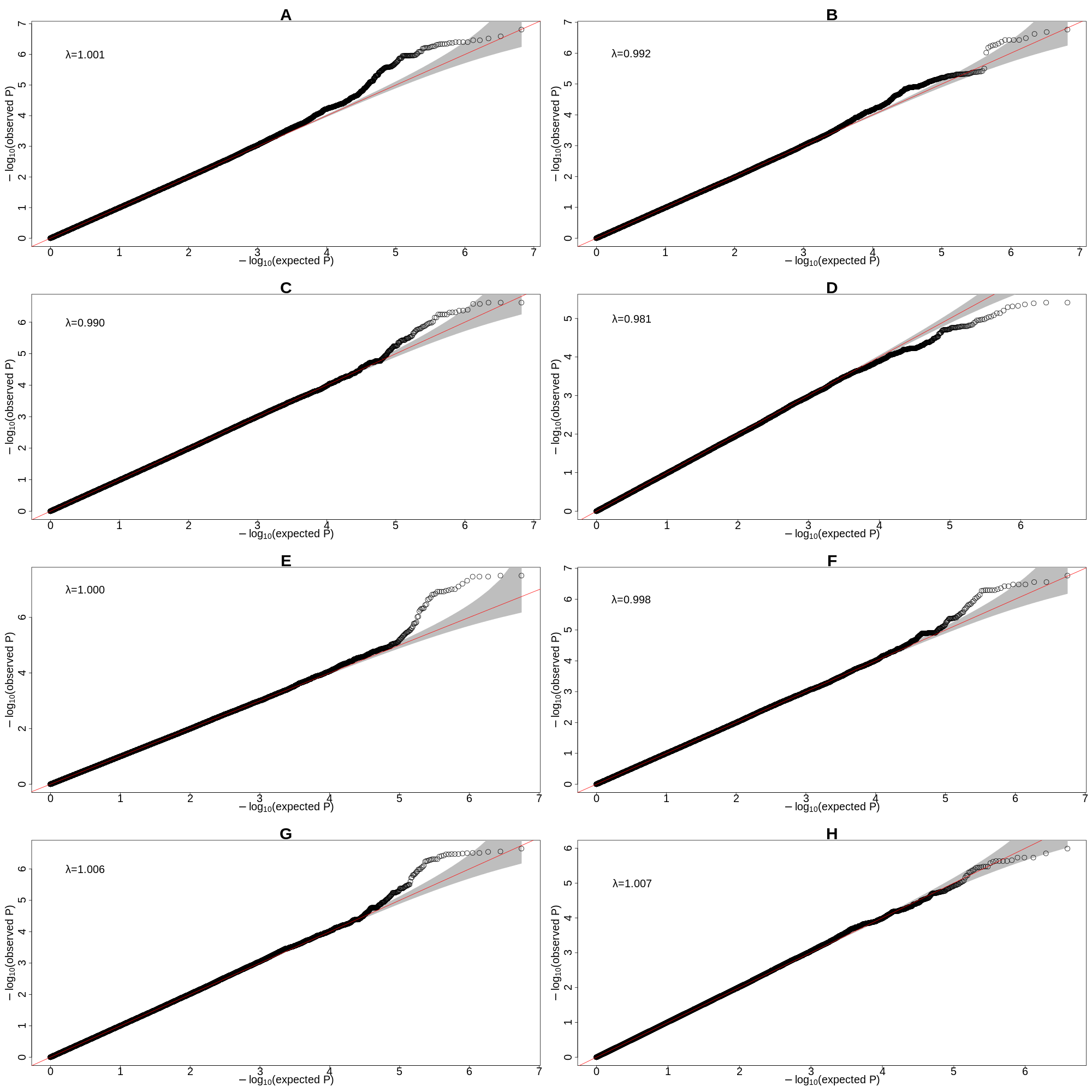

**Supplemental Figure 2. Quantile-Quantile (QQ) plots from each TRACTOR and META GWAS performed. A.** Tractor GWAS African Tract, no conditioning **B.** TRACTOR GWAS African Tract, conditioned on *VKORC1*-1639G>A (rs9923231) **C.** TRACTOR GWAS African Tract, conditioned on VKORC1-1639G>A (rs9923231), CYP2C9*2, and CYP2C9*3 **D.** TRACTOR GWAS African Tract, conditioned on *VKORC1*-1639G>A (rs9923231), *CYP2C9**2, *CYP2C9**3, *CYP2C9**8, and *CYP2C9**11 **E.** META GWAS, no conditioning **F.** M META GWAS, conditioned on *VKORC1*-1639G>A (rs9923231) **G** META GWAS , conditioned on *VKORC1*-1639G>A (rs9923231), *CYP2C9**2, and *CYP2C9**3 **H.** META GWAS , conditioned on *VKORC1*-1639G>A (rs9923231), *CYP2C9**2, *CYP2C9**3, *CYP2C9**8, and *CYP2C9**11

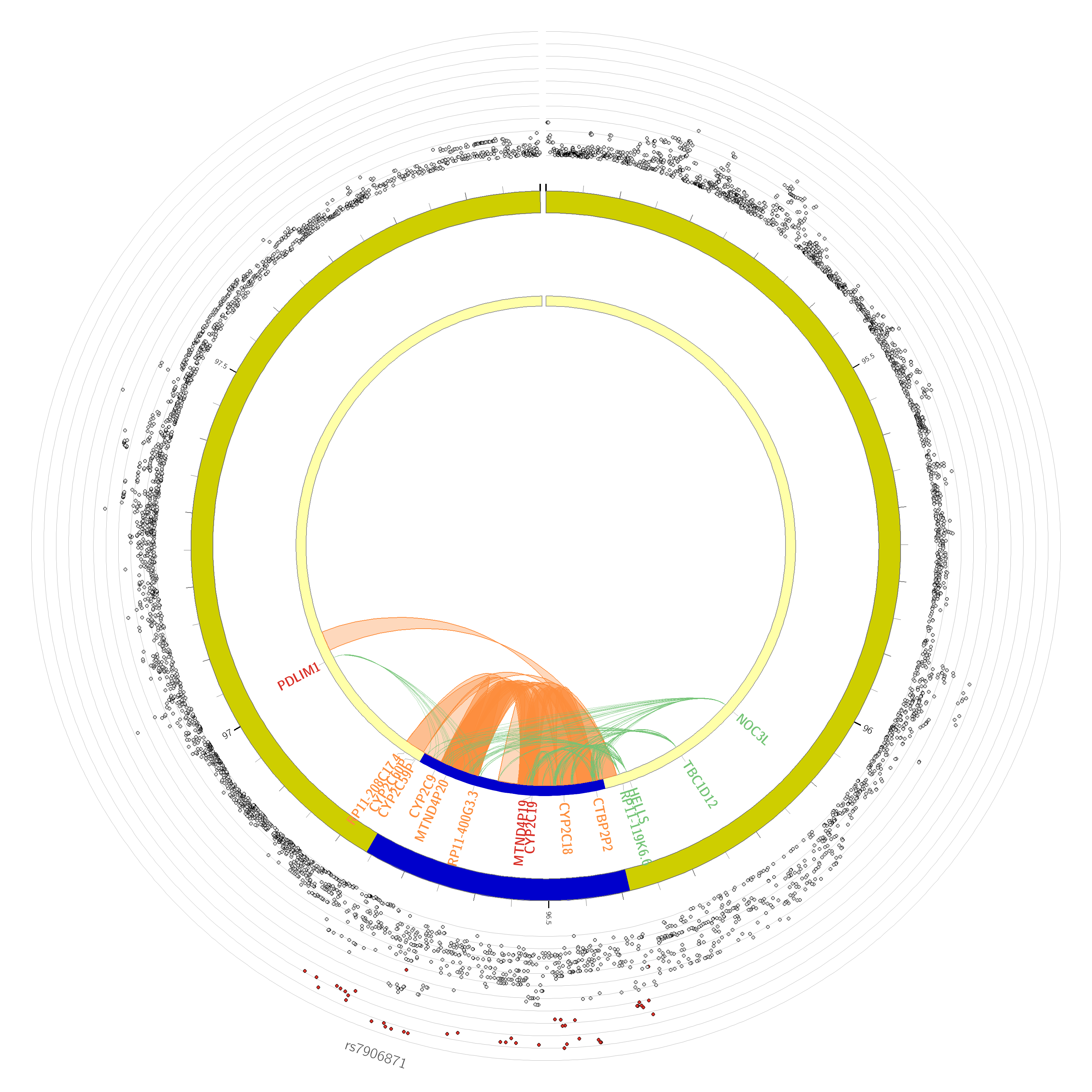
**Supplemental Figure 3. Chromatin Interaction Plot from FUMA [6].** Shows

indication that rs7906871 is involved in several highly significant chromatin interactions within the CYP2C locus (FDR = 4.52E-7 – 3.59E-23). The outermost layer of the plot is a Manhattan plot of the SNPs located within chromosome 10 with SNPs colored based on the LD with rs7906871 (red (r^2^ > 0.8), orange (r^2^ > 0.6), green (r^2^ > 0.4), blue (r^2^ > 0.2), and grey (r^2^≤ 0.2). The middle and innermost rings both represent chromatin interactions with the region highlighted in blue representing the genomic risk loci within chromosome 10. The orange links within the innermost ring represent the chromatin interactions.

**Supplemental Tables Legends**

**Supplemental Table 1. Pearson Correlation Test Results for potential covariates to include in TRACTOR GWAS.** Each covariate that was pulled from the IWPC cohort was tested against stable warfarin dose for Pearson's correlation by using the cor.test() function in R version 4.4.0. Variables that had a significant correlation estimate with warfarin dose were used as covariates in the TRACTOR GWAS.

**Supplemental Table 2. SNPs that met the suggestive significance threshold (P<5E-6) in the TRACTOR African tract for the TRACTOR GWAS in the IWPC Cohort (consists of 340 AAs on stable warfarin dose).** 128 SNPs met this threshold in the TRACTOR African tract and were included in this table. The Effect Allele frequency, the frequency of the allele that was tested for association by TRACTOR, as well as the Beta in the African tract for each of these SNPs is included.

**Supplemental Table 3.** **SNPs that met the suggestive significance threshold (P<5E-6) in the IWPC AA cohort (N=340) and were replicated (P<1E-3) with the same direction of effect in the ACCOuNT cohort (N=309).** 35 SNPs that met the suggestive significance threshold in the IWPC cohort were replicated in the ACCONT cohort which adds more weight to the association of these SNPs to stable warfarin dose.

**Supplemental Table 4. SNPs that met the suggestive significance threshold (P<5E-6) in the Ancestry Tract Meta-Analysis in the IWPC Cohort (consists of 340 AAs on stable warfarin dose).** 49 SNPs met the suggestive significance threshold in the ancestry tract meta-analysis and were included in this table. The Effect Allele frequency, the frequency of the allele that was tested for association, as well as the meta-analyzed beta for each of these SNPs is included.

**Supplemental Table 5. SNPs that met the nominal significance threshold (P<1E-4) in the Ancestry Tract Meta-Analysis conditioned on *VKORC1*-1639G>A (rs9923231) in the IWPC Cohort (consists of 340 AAs on stable warfarin dose).** 453 SNPs still retained significance after adjustment for rs9923231, including the lead SNP in this analysis rs7906871 (P=3.33E-7). The Effect Allele frequency, the frequency of the allele that was tested for association, as well as the meta-analyzed beta for each of these SNPs is included.

**Supplemental Table 6. SNPs that met the met the nominal significance threshold (P<1E-4) in the Ancestry Tract Meta-Analysis conditioned on *VKORC1*-1639G>A (rs9923231) and *CYP2C9* *2 and *CYP2C9**3 variants in the IWPC Cohort (consists of 340 AAs on stable warfarin dose).** 506 SNPs still retained significance after adjustment for rs9923231 and *CYP2C9* (*2 and *3), including the lead SNP in this analysis rs7906871 (P=3.35E-7). The Effect Allele frequency, the frequency of the allele that was tested for association, as well as the meta-analyzed beta for each of these SNPs is included.

**Supplemental Table 7. SNPs that met the met the nominal significance threshold (P<1E-4) in the Ancestry Tract Meta-Analysis conditioned on *VKORC1*-1639G>A (rs9923231) and *CYP2C9* locus (*2, *3, *8, and *11 variants) in the IWPC Cohort (consists of 340 AAs on stable warfarin dose).** 319 SNPs still retained significance after adjustment for rs9923231 and *CYP2C9* locus, including the lead SNP in this analysis rs7906871 (P=9.40E-5). The Effect Allele frequency, the frequency of the allele that was tested for association, as well as the meta-analyzed beta for each of these SNPs is included.

**Supplemental Table 8. SNPs that met the suggestive significance threshold (P<5E-6) in the GWAS done in the IWPC European Cohort (consists of 1259 Europeans on stable warfarin dose).** 21,751 SNPs met this threshold in the IWPC Europeans and were included in this table. The effect allele frequency as well as the Beta for each of these SNPs is included. The lead SNP in the TRACTOR analysis, rs7906871, met the suggestive significance threshold in the IWPC European cohort as well (P=4.68E-6) indicating that it may be associated to stable warfarin dose in Europeans and AAs.

**Supplemental Table 9. SNPs that met the nominal significance threshold (P<1E-4) in the GWAS conditioned on *VKORC1*-1639G>A (rs9923231) done in the IWPC European Cohort (consists of 1259 Europeans on stable warfarin dose).** 35,266 SNPs met this threshold after conditioning on rs9923231 in the IWPC European cohort. The effect allele frequency as well as the Beta for each of these SNPs is included. The lead SNP in the TRACTOR analysis, rs7906871, is still nominally significant after conditioning on rs9923231 in the IWPC European cohort (P=1.43E-5) indicating that its effect on stable warfarin dose is independent of rs9923231 in Europeans and AAs.

**Supplemental Table 10. SNPs that met the nominal significance threshold (P<1E-4) in the GWAS conditioned on *VKORC1*-1639G>A (rs9923231), *CYP2C9**2, and *CYP2C9**3 done in the IWPC European Cohort (consists of 1259 Europeans on stable warfarin dose).** 34,585 SNPs met this threshold after conditioning on rs9923231, *CYP2C9**2, and *CYP2C9**3 in the IWPC European cohort. The effect allele frequency as well as the Beta for each of these SNPs is included. The lead SNP in the TRACTOR analysis, rs7906871, is not significant after adding *CYP2C9**2 and *CYP2C9**3 in the IWPC European cohort and thus was not included in this table (P=0.12), indicating that in Europeans its signal is absorbed by the *CYP2C9* star alleles.

**Supplemental Table 11. Genomic Loci prioritized by FUMA [5] with independent significant SNPs and Lead SNPs listed within each cluster.** Table that shows the 13 genomic risk loci prioritized by FUMA [5] as well as the location of the independent lead SNP and rsID’s of independently significant SNPs (Significant GWAS SNPs that have a r^2^<0.8 with each other) located within each cluster.
